# Compound risks of hurricane evacuation amid the COVID-19 pandemic in the United States

**DOI:** 10.1101/2020.08.07.20170555

**Authors:** Sen Pei, Kristina A. Dahl, Teresa K. Yamana, Rachel Licker, Jeffrey Shaman

**Author notes:** These authors contributed equally to this article (Sen Pei, Kristina A. Dahl).

## Abstract

Current projections and unprecedented storm activity to date suggest the 2020 Atlantic hurricane season will be extremely active and that a major hurricane could make landfall during the global COVID-19 pandemic. Such an event would necessitate a large-scale evacuation, with implications for the trajectory of the pandemic. Here we model how a hypothetical hurricane evacuation from four counties in southeast Florida would affect COVID-19 case levels. We find that hurricane evacuation increases the total number of COVID-19 cases in both origin and destination locations; however, if transmission rates in destination counties can be kept from rising during evacuation, excess evacuation-induced case numbers can be minimized by directing evacuees to counties experiencing lower COVID-19 transmission rates. Ultimately, the number of excess COVID-19 cases produced by the evacuation depends on the ability of destination counties to meet evacuee needs while minimizing virus exposure through public health directives.

## Introduction

The combination of the COVID-19 pandemic, existing racial and socioeconomic inequalities, and environmental stressors exacerbated by climate change is exposing the many ways in which “compound risks” threaten human lives and wellbeing while straining the ability of governments at all scales to limit the damage from any one threat on its own^1^. Intersections of climate extremes with the pandemic—recent widespread flooding in South Asia at a time of rapidly increasing COVID-19 caseloads, for example— have made clear that the consequences of such compound risk events can be lethal, though the underreporting of cases around the world^2^ and widely varying testing capabilities^3^ make it difficult to accurately quantify their magnitude.

With the anticipated peak of the 2020 Atlantic hurricane season approaching and COVID-19 cases widespread and abundant in many hurricane-prone areas of the United States, the nation is poised to experience the collision of two major disasters. This study therefore addresses how decision-making around one key aspect of hurricane response—evacuation—could influence the trajectory of the pandemic in the US and be optimized to limit excess COVID-19 cases. With future global warming expected to continue the observed trend toward increasingly intense Atlantic hurricanes^4,5^, understanding how to manage and minimize the impact of the combined risks associated with a major hurricane and a global pandemic could prove critical both later this year and in the long-term as the risks of such simultaneous disasters increase around the world.

Efficient, effective evacuations—whether voluntary or mandatory—are a critical component of ensuring public safety during natural disasters. The scale of recent evacuations from US Southeast and Gulf Coast states has been large: During Hurricanes Matthew (2016), Irma (2017), and Dorian (2019), for example, roughly 2.5, 6.5, and 1.1 million people, respectively, were under evacuation orders ^6-8^. By changing the distribution of people for days or weeks, a large-scale evacuation amid a pandemic has the potential to alter the trajectory and geographic distribution of infections. And by temporarily relocating from their own homes into potentially shared living arrangements where levels of social contact are higher, evacuees may experience greater transmission risk both during and after an evacuation.

This analysis evaluates how a large-scale evacuation of the southeast Florida coast from a hypothetical Category 3 hurricane would affect the total number of COVID-19 cases and their spatial distribution in evacuees’ origin and destination counties. This is accomplished by first building a hypothetical hurricane evacuation scenario from previously published studies of evacuation behavior in the southeast US region. We then use a simple, two-county infectious disease model to identify the most relevant evacuation and epidemiological characteristics influencing COVID-19 case counts. The findings from the these simulations are used to inform experiments with a larger metapopulation model representing SARS-CoV-2 transmission in all 3,142 US counties^9^, as well as various evacuation scenarios.

## RESULTS

### Identifying key parameters using a two-county model

We used a simplified, two-county metapopulation model, representing a generic pair of origin and destination counties, to determine the factors that have greatest influence on COVID-19 case numbers (Methods; Figure 1a). Specifically, we evaluated the effects of six evacuation and epidemiological characteristics on COVID-19 case numbers: transmission rates in origin and destination counties (quantified by the effective reproductive number, *R_e_*), the fraction of the origin county population that evacuates (*p_eva_*), the duration of the evacuation period (*T_eva_*), and daily case numbers in the origin and destination counties (*case_ori_* and *case_dest_*). We simulated an evacuation by moving a fraction of the population from the origin to the destination county. Evacuees then mixed with the population of the destination county, before returning home. This simulation was repeated using different combinations of each of the six characteristics in order to determine the effects of each on COVID-19 case numbers during and following the evacuation.

**Figure 1:**
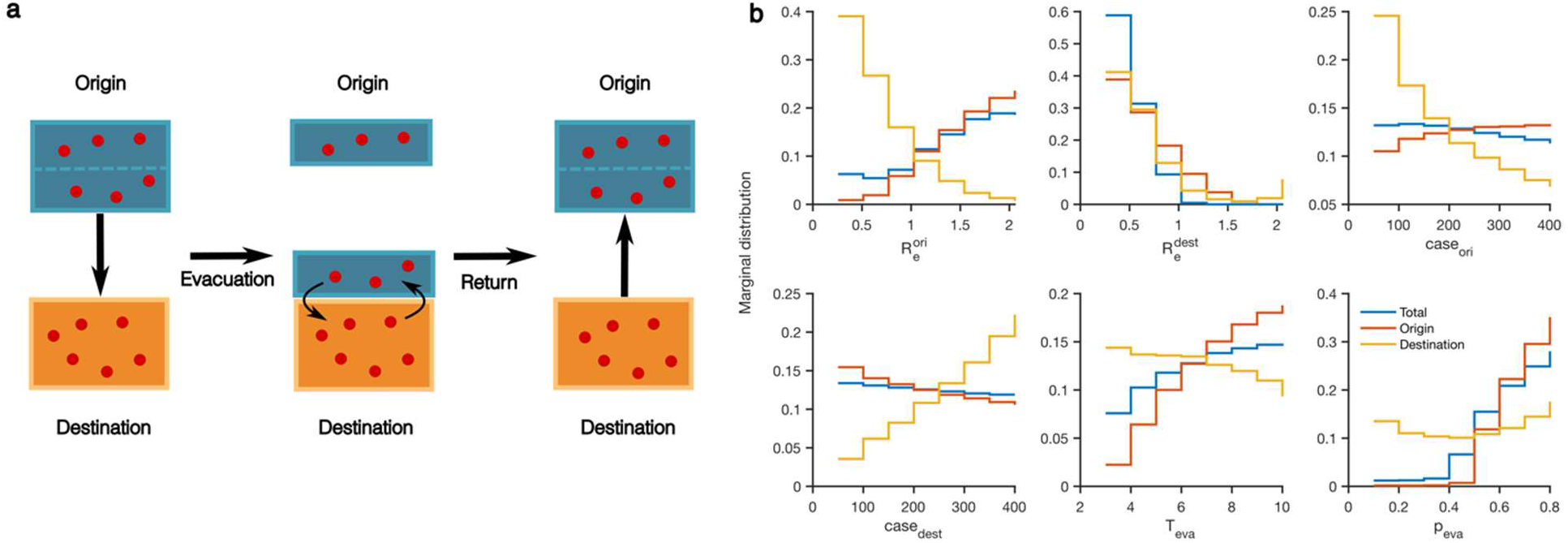
Results from the two-county model showing that origin and destination transmission rates have the greatest influence on final case numbers. (a) A schematic diagram for the two-county model. Blue and orange boxes represent the origin and destination populations. Red dots within boxes represent infected individuals. (b) The marginal distribution of six parameters for the top 10% of combinations that lead to the lowest percentage increase (or highest percentage reduction) of reported cases in the origin county (solid red lines), the destination county (solid orange lines) and both counties combined (solid blue lines). Here 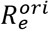 and 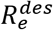 represent the transmission rates in the origin and destination; *case_ori_* and *case_dest_* represent the daily cases in the origin and destination; *T_eva_* is the duration of evacuation; and *p_eva_* is the fraction of the origin population evacuating.

We found that transmission rates in the origin and destination counties were the primary determinant of case numbers (Methods; Figure 1b): evacuating individuals from a high-*R_e_* origin to a low-*R_e_* destination produced fewer additional cases in the origin county and in the origin and destination county combined. For the destination county alone, it was preferable to accept evacuees from a low-*R_e_* origin. However, in a real hurricane landing, the counties that require evacuation are determined by the path of the hurricane, i.e. a low-*R_e_* origin cannot be stipulated. The length of evacuation and the number of people evacuating also influenced case numbers; however, these two characteristics are also expected to be shaped more by the specific circumstances necessitated by a particular hurricane rather than by public health directives.

### Full model simulations of hurricane evacuation scenarios

Next we used a national county-scale metapopulation model and a suite of scenarios to further explore how transmission rates and hurricane evacuation affect COVID-19 incidence in origin and destination counties (Methods). All scenarios assume that a Category 3 hurricane is approaching southeast Florida and that people living in Palm Beach, Broward, Miami-Dade, and Monroe Counties are ordered to evacuate. Based on studies of evacuation compliance and behavior in this region for Category 3+ hurricanes, we estimate that 48% of each county’s population would evacuate^7,10-17^ (Methods). Assuming that 19% of evacuees relocate elsewhere within their respective counties, this leads to a total of 2.3 million evacuees leaving the four affected counties^7,16-20^.

For each scenario, evacuees were assigned to a different set of destination counties based on a list of 165 possible destinations across 26 states identified during post-Hurricane Irma surveys^7,17^. In the baseline scenario, evacuees were assigned to all 165 destination counties in proportion to observed evacuation choices during Hurricane Irma. The number of evacuees assigned to each destination county for this baseline scenario is shown in Figure 2a. In order to explore the effects of destination county transmission levels on the number of evacuation-associated COVID-19 cases, we further proposed two hypothetical scenarios in which evacuees were assigned to locations with high *R_e_* or low *R_e_*. In the high (low) *R_e_* scenario, 90% of the evacuees assigned to each county in the baseline scenario were instead diverted to the 82 counties with the highest (lowest) *R_e_*, weighted by the proportion of evacuees sent to each of these counties in the baseline scenario (Methods).

**Figure 2:**
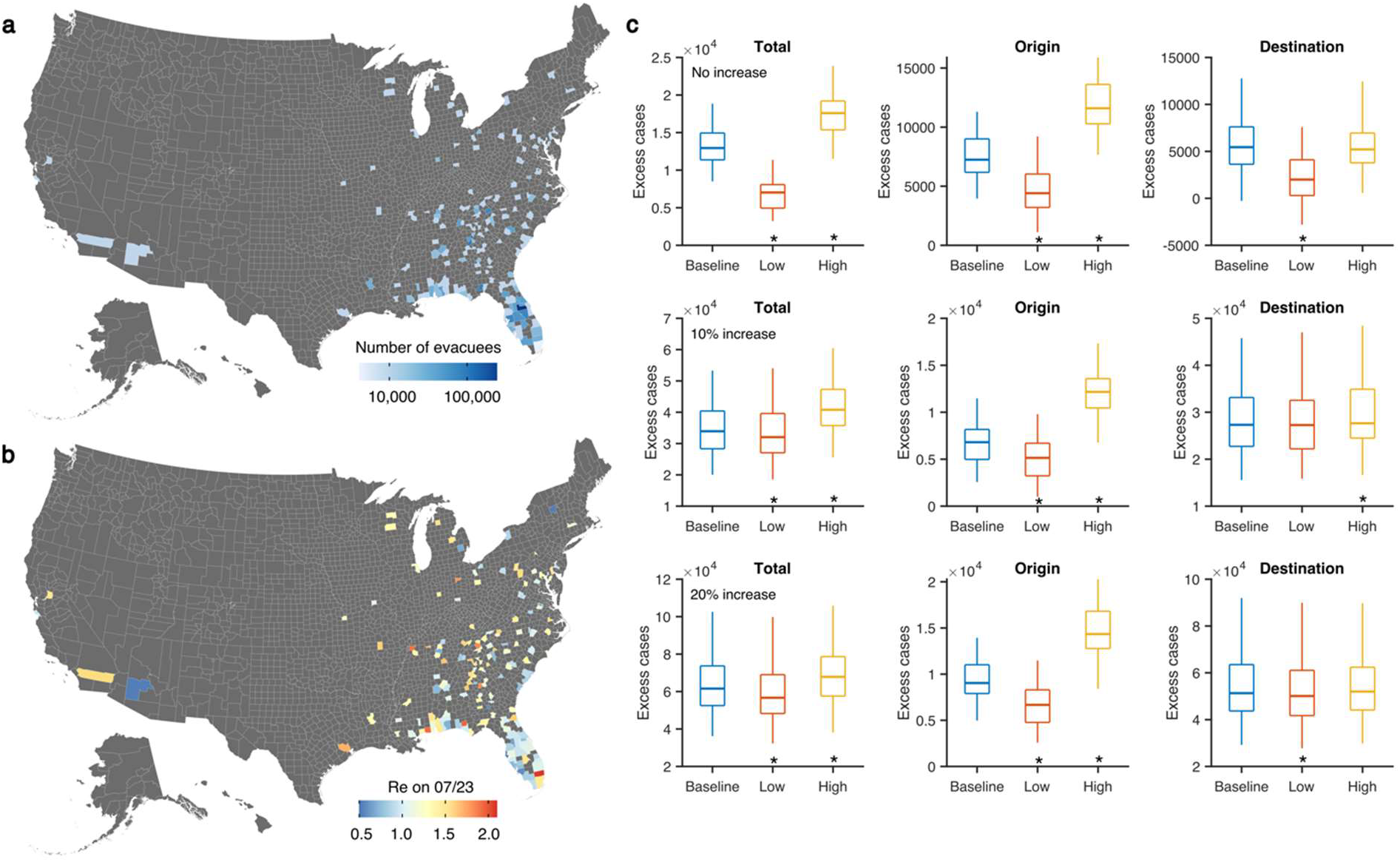
Simulations for evacuation using the national county-level transmission model. (a) The number of evacuees accepted by 165 destination counties in the baseline scenario. (b) The estimated effective reproductive numbers *R_e_* for both origin and destination counties on July 23^rd^, 2020. (c) Comparison of excess cases in origin and destination counties combined (left column), only origin counties (middle column) and only destination counties (right column) for the baseline, low and high evacuation scenarios. Simulations were performed for three settings: no increase (top row), 10% increase (middle row) and 20% increase (bottom row) of the transmission rates in destination counties. Box plots show the median and interquartile and whiskers show the 95% CIs. Asterisks indicate that excess cases are significantly lower or higher than the baseline scenario (Wilcoxon signed rank test, *p <* 10^−5^). Results are obtained from 100 model simulations.

Scenario projections were initiated from the model state calibrated to observed county-level COVID-19 case and death data from February 21^st^ through July 23^rd^, 2020 (Methods). The estimated effective reproductive numbers (*R_e_*) in origin and destination counties at the start of simulations are shown in Figure 2b. Evacuees tend to stay with friends or family, in hotels/motels, or in public shelters, each of which would likely increase transmission opportunities relative to simply staying home^7^. To reflect this, we assume the COVID-19 transmission rate in destination counties increases during the evacuation period by either 0%, 10%, or 20%. These settings implicitly differentiate the levels of control effected in destinations during evacuation, as well as differences in transmission potential associated with different types of accommodation (e.g., staying with friends/families, hotels or shelters). In addition, to reflect periods of hurricane preparation and recovery^21-23^, we elevated the transmission rate in the origin counties by 20% beginning 3 days prior to evacuation and ending 3 days after the return of evacuees (more detailed simulation settings are provided in Methods). For comparison, we also generated simulations for the same period but without evacuation.

In all scenarios, combined cases in origins and destinations are primarily driven by ongoing local COVID-19 transmission dynamics (Figure S1; Table S1); however, evacuation does alter disease outcomes. In the baseline scenario, total COVID-19 cases in the origin and destination counties increase significantly relative to the no-evacuation scenario (Figure 2c; Table 1; Wilcoxon signed rank test), indicating evacuation in and of itself can cause a statistically significant increase of COVID-19 cases. As indicated by the low and high scenarios, the number of COVID-19 cases resulting from evacuation is significantly lower (higher) than the baseline scenario if evacuees are directed to counties with lower (higher) transmission rates. However, as the transmission rate in the destination counties increases, the differences between the low and high scenarios become less pronounced. This result indicates that the benefits of a directed evacuation would be amplified by more stringent control efforts in destination counties.

**Table 1:** Full metapopulation model simulation of the median number of excess cases in origin and destination counties for different evacuation scenarios (baseline, low, high, and optimized) and different increases of transmission rates (*R_e_*) in destination counties (no change, 10% increase, 20% increase). Note that the high and low *R_e_* scenarios are not subject to the constraint of destination capacity, whereas the optimized scenario takes into account a hypothetical capacity for each destination county.

| | 0% $R_e$ increase in destination | | 10% $R_e$ increase in destination | | 20% $R_e$ increase in destination | |
| --- | --- | --- | --- | --- | --- | --- |
|  | Origin excess cases | Destination excess cases | Origin excess cases | Destination excess cases | Origin excess cases | Destination excess cases |
| **Baseline evacuation** | 7,244 | 5,448 | 7,853 | 28,661 | 8,973 | 52,478 |
| **High $R_e$ evacuation** | 11,593 | 5,209 | 12,173 | 27,649 | 14,338 | 51,996 |
| **Low $R_e$ evacuation** | 4,409 | 1,999 | 5,143 | 27,270 | 6,669 | 50,080 |
| **Optimized evacuation** | 5,441 | 3,628 | 4,989 | 25,919 | 7,333 | 50,724 |

### Greedy optimization method to minimize COVID-19 cases

The model simulations for our hypothetical evacuation scenarios indicate that a strategic evacuation plan could reduce excess COVID-19 infections. However, these scenarios neither accounted for the accommodation capacities of destination counties nor provided a framework for optimizing evacuation plans.

To address these issues, we developed a greedy search optimization algorithm aimed at minimizing total excess COVID-19 cases by strategically assigning evacuees to optimal destination counties. As indicated by the two-county model simulations, evacuating individuals to destinations with low *R_e_* reduces COVID-19 transmission. However, given the varying prevalence of infection in origin counties and the nonlinear transmission dynamics, it is not straightforward to determine the optimal number of evacuees from each origin county that should be prioritized and redirected to each of the lowest-*R_e_* destinations. In conducting the optimization, we imposed the following constraints on human movement: 1) we assumed that a fraction of evacuees cannot be redirected from their baseline destination county, representing individuals whose choice of destination will not be influenced by evacuation directives, perhaps due to financial constraints or preferences to stay with family; and 2) we prescribed a capacity limit on the number of evacuees received by each destination county. The greedy search starts from an evacuation matrix representing the evacuees who cannot be redirected from their destination and then iteratively directs the remaining evacuees to destination counties with lowest *R_e_*. In each iteration, the algorithm selects which origin counties will be assigned the evacuee slots available in a destination county. This search is repeated for each successive destination county until all evacuees are assigned a destination (Methods, Supplementary Information).

We repeated this evacuation optimization with three different settings: no increase, 10% increase, and 20% increase of transmission rates in destination counties, again to reflect differences in control efforts and accommodation type. We assumed that 10% of evacuees will maintain their original destination and are thus not redirected, and that each destination has a capacity of 120% of the evacuees accepted in the baseline scenario. We then generated the optimized evacuation plan for each setting. During the optimization process, assigning more evacuees to low-*R_e_* counties led to a reduced number of total infections compared to the baseline scenario (Figure S2). The optimized top 20 destinations for the four origin counties are reported in Tables S2-S4. In Figure 3a, we show the change in the number of evacuees to each destination for the optimized evacuation scenario with a 10% increased transmission rate in destination counties. In general, evacuees who traveled to high-*R_e_* destinations in the baseline scenario were redirected to low-*R_e_* destinations. For all three transmission rate scenarios, the optimized evacuation effectively reduces the number of excess cases in both origin and destination counties compared to the baseline scenario (Figure 3b). The reduction is greatest (up to 30%) for the scenario in which there is no increase in the transmission rate in destination counties, which highlights the crucial role of effective intervention during evacuation. In this optimization example, the fraction of non-allocable evacuees and destination capacity are hypothetical, as these quantities are unknown. If such information were available or could be estimated using socioeconomic and geographic characteristics, the optimization could be tailored to reflect more realistic constraints on evacuation.

**Figure 3:**
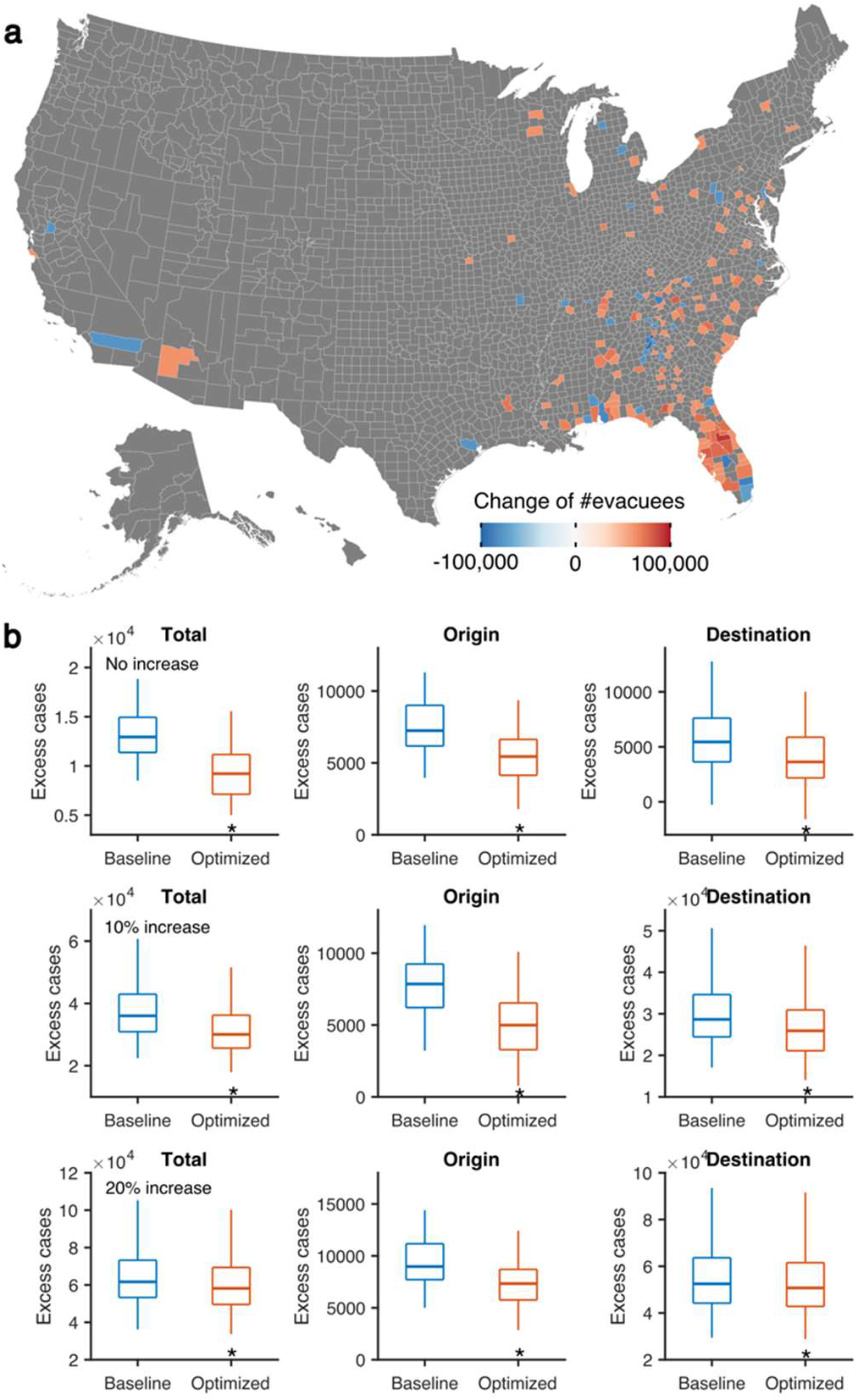
Optimization of evacuation plans. (a) The change in the number of evacuees to destination counties in the optimized evacuation plan compared with the baseline evacuation scenario. Evacuation was optimized for the setting in which transmission rates in destination counties increase by 10%. (b) Excess cases for the baseline and optimized evacuation scenarios are compared for the origin and destination counties combined (left column), only origin counties (middle column) and only destination counties (right column). Simulations were performed for three settings: no increase (top row), 10% increase (middle row) and 20% increase (bottom row) of the transmission rates in destination counties. Boxes and whiskers show the median, interquartile and 95% CIs. Asterisks indicate that excess cases are significantly lower than the baseline scenario (Wilcoxon signed rank test, *p <* 10^−5^). Results are obtained from 100 model simulations.

## DISCUSSION

The results of this study have far-reaching consequences not only for hurricane evacuation this season but also for long-term US hurricane preparedness and evacuation planning.

Research suggests that people rely on past experiences when choosing their evacuation routes and destinations^20,24^. This study shows that excess COVID-19 cases could be minimized by instead directing evacuees to either counties with lower COVID-19 transmission rates or an optimized set of counties. While decisions about whether to evacuate and where to go ultimately fall to individual households, emergency communications from federal agencies and broadcast meteorologists can influence residents’ perceptions of hurricane threats and are seen as trusted sources of information in emergency situations^25^. Because a majority of US residents are concerned about COVID-19^26^, if the need for a large-scale evacuation arises, evacuees may turn to these same trusted sources for information on how best to stay safe while evacuating. Local, state, and federal officials who develop evacuation orders and communicate them to the general public may therefore want to consider whether their evacuation-related communications should include assessments of the relative safety of potential destination counties with respect to COVID-19 risk rather than allowing default evacuation patterns based on past storms to prevail.

This research shows that the magnitude of the impact of evacuation on COVID-19 caseloads is highly dependent on conditions in destination counties. The degree to which counties are prepared to host, isolate, and meet the needs of evacuees while also minimizing virus exposure through public health directives such as social distancing and mask wearing will be a key determinant of the impact of evacuation on COVID-19 case numbers. Preparedness within destination counties is particularly important because, as this analysis shows, destination counties will bear the brunt of the excess COVID-19 cases that result from an evacuation event. Destination counties must be aware of the influx of evacuees should it occur and must be allocated the financial and human resources needed to ensure the safety of both their residents and the evacuees they are sheltering.

The US response to the COVID-19 pandemic has varied widely from state to state and from county to county. As a result of policy, communication and ideological differences, compliance with mask wearing, for example, has varied substantially even within a given state^27^. This variability could extend into county-level hurricane preparedness measures, particularly given that guidance may be issued at the state level while implementation of specific measures is left to counties, as is the case for Florida’s current co-response guidance on hurricane evacuation and COVID-19^28^.

Centuries of systemic racism in the US have left Black, Native American, Latinx, and other non-White people with both higher exposure to and fewer resources to cope with environmental or health-related stressors compared with White populations^29,30^. For example, recent research suggests that federal financial aid after natural disasters is not equitably distributed among communities and may even exacerbate income inequality^31,32^. Low-income communities and communities of color consequently struggle to prepare in advance of and recover in the wake of natural disasters^33,34^.

Due to systemic health inequities, including higher exposure to air pollution^29^ and higher rates of underlying health conditions^35^, COVID-19 has also disproportionately affected Black, Native American, and Latinx people in the US^36^. These groups have experienced higher infection rates, poorer health outcomes, and deeper declines in employment during the pandemic^36-38^. The additional risks faced because of COVID-19 and the financial costs associated with evacuation^39^ could present additional challenges for these segments of the population during hurricane evacuation or discourage them from evacuating altogether.

This study has used idealized scenarios to model hurricane evacuation patterns. These scenarios cannot fully capture many household-level choices that could alter levels of social contact—and therefore potential COVID-19 exposure—during the evacuation period. For example, previous studies of hurricane evacuations along the US Southeast and Gulf Coasts show that evacuees strongly and consistently prefer to stay with friends and family over going to hotels/motels or public shelters^7,14 15,20,39,40^. Levels of social contact and potential virus transmission would likely differ across accommodation types, which implies a level of complexity and spatial heterogeneity that is not possible to incorporate within the model used in this study. Similarly, this study does not consider variable levels of exposure to COVID-19 based on evacuation transportation mode. While evacuees strongly prefer to travel in their own vehicles^17^, shared modes of transportation such as buses or carpools would increase potential virus transmission and exposure. Recent research suggests that for both transportation and shelter, the sharing economy— Internet-based transactions via companies like Airbnb that allow for peer-to-peer sharing of goods and services—could play a role in providing free or affordable resources to evacuees that would enable evacuees to maintain social distancing^41^.

As a result of the ongoing economic and physical toll of the pandemic, household-level decision making regarding evacuation may differ from that of past years. There are many sociodemographic factors associated with decision making around evacuation including experience with past hurricanes, length of residence, home ownership, age, income, race, employment status, level of social connectivity, social cues, perceived levels of self-efficacy and risk, and storm conditions^42-46^. While some of these factors (e.g. gender and race) are unchanged from last year, others (e.g. employment status and income) may be either changed or very much influenced by the current COVID-19 pandemic. In contrast, the idealized scenarios adopted for this study assume that people will choose destination counties and accommodation types that match past choices.

The movement of people in and out of hurricane-affected counties does not simply cease after all evacuees have returned to their homes. For instance, communities affected by hurricanes often experience an influx of workers who assist with rebuilding and recovery efforts^47-49^, which could also influence infection rates in the affected counties long after the evacuation period. Post-evacuation movement patterns are beyond the scope of the present study.

Critically, hurricane evacuation is intended to save lives and prevent serious injuries to residents of hurricane-prone regions. While this study evaluates excess COVID-19 cases resulting from evacuation, it does not evaluate non-COVID-19 related risks to human health and lives in the event that people choose to remain in their homes despite receiving evacuation orders-risks that could increase if people are afraid to evacuate out of concern for contracting COVID-19. Nor does it address evacuations of hospitals, nursing homes, prisons, or other facilities. It will be critical for emergency managers to factor in these— and other—complexities when developing plans^50^.

Finally, the results presented here are based on scenarios that, while plausible, are strictly hypothetical. While the overall notion that distributing evacuees to destination counties with low transmission rates minimizes excess cases should theoretically apply to geographies outside Florida or the US, additional model simulations of such scenarios should be generated.

## CONCLUSION

The data presented here show that while a large-scale hurricane evacuation would increase the total number of COVID-19 cases in the US, directing evacuees to plausible destination counties with low COVID-19 transmission rates would minimize the excess cases induced by the evacuation event. These results have far-reaching implications for immediate emergency management and communications practices, as well as long-term disaster preparedness.

Faced with the prospect of tens of thousands of additional cases arising from a hurricane evacuation, states and counties at both ends of evacuation routes must be allocated the necessary financial and human resources required to meet evacuees’ needs while also ensuring community safety and health through measures intended to reduce COVID-19 transmission rates. Further, resource distribution must prioritize the nation’s most vulnerable groups.

## Data Availability

Data and code are available on github.

https://github.com/SenPei-CU/Hurricane-COVID

## ACKNOWLEDGEMENTS

The authors would like to acknowledge Stephen Wong, who graciously provided data that informed this analysis. Astrid Caldas, Rachel Cleetus, Juan Declet-Barreto, Adrienne Hollis, and Erika Spanger-Siegfried provided inspiration and insight during the course of the research. Work on this study was supported by NSF grant DMS-2027369, a gift from the Morris-Singer Foundation, and the generous support of the Barr Foundation, the Common Sense Fund, the Energy Foundation, Farvue Foundation, the Fresh Sound Foundation, the MacArthur Foundation, New York Community Trust, the Rauch Foundation, Sand County Charitable Trust, The Scherman Foundation, three anonymous funders, and members of the Union of Concerned Scientists.

## METHODS

### Hurricane evacuation scenario development

To develop a hypothetical hurricane evacuation scenario for southeast Florida, we drew from previously published hurricane evacuation studies focused on Category 3+ hurricanes that have affected the Southeast or Gulf Coast regions of the US. In this scenario, we assumed a Category 3 hurricane approaching the southeast Florida coast along a track that would necessitate evacuations from Palm Beach^51^, Broward^52^, Miami-Dade^53^, and Monroe^54^ Counties.

We obtained the population under mandatory evacuation orders in each county from GIS shapefiles of the zones of mandatory evacuation from a Category 3+ hurricane for each county^55^. We then calculated the percent of the population living within mandatory evacuation zones that would actually comply with evacuation orders by averaging the compliance rate from eight regionally relevant studies of evacuation behavior^7,10-16^. We found that the average evacuation order compliance rate from these studies was 66%. Because many people living outside of the mandatory evacuation zones voluntarily choose to evacuate during hurricanes as well, we used the same approach to calculate the percent of the population living outside of mandatory evacuation zones but within the affected counties that would evacuate. Based on four studies of this “shadow evacuation” phenomenon, we determined that an average of 47% of county residents outside mandatory evacuation zones would also evacuate^7,13-15,17^. The mandatory and voluntary evacuees together represent 48% of each origin county’s population. We then determined that 19% of these evacuees would relocate within their respective counties based on the average from four evacuation behavior studies^7,16-20^.

Finally, to determine the destination counties of evacuees from each of the four origin counties, we obtained raw post-Hurricane Irma survey data^7,17^. These data allowed us to identify both the destination counties and the percent of evacuees choosing each destination county. We then apportioned evacuees leaving the four origin counties to each destination county.

### Two-county model of COVID-19 transmission

In order to identify the most sensitive factors driving the COVID-19 transmission during evacuation, we first ran simulations using a simple two-county model. This model describes the transmission dynamics of COVID-19 during a hurricane evacuation from an origin (county 1) to a destination (county 2). Mathematically, the transmission dynamics in the origin and destination are depicted by a susceptible-exposed-infected-recovered (SEIR) model. We simulate the disease transmission as a stochastic Markov process using the following equations:

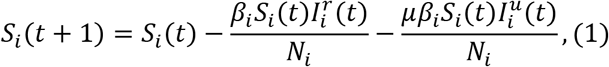

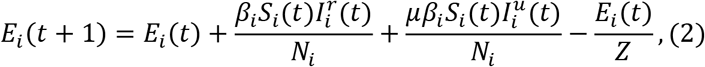

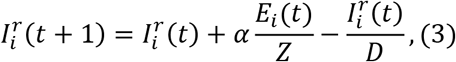

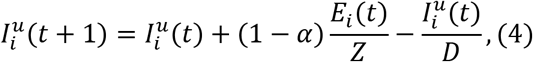

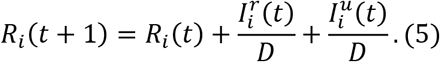

Here *N_i_*, *S_i_*(*t*)*, E_i_*(*t*), 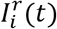, 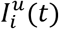 and *R_i_*(*t*) are the total, susceptible, exposed, reported infected, unreported infected and recovered population in county *i* on day *t; β_i_* is the transmission rate in county *i*; *μ* is the relative transmissibility for unreported infections; *Z* is the average latency period; *D* is the average duration of infectiousness; *α* is the fraction of reported infections. The effective reproductive number, which quantifies the local transmission rate, is computed as *R_e_ = β_i_D[α + μ*(1 − *α*)*]S_i_/N_i_* using the next generation matrix approach.

During evacuation, we assume a fraction (*p_eva_*) of the population is evacuated from county 1 to county 2 and mixes with the local population for *T_eva_* days. Individuals within each compartment are randomly drawn from the population in the origin. We track the infections in the evacuated population in county 2, which then return to the origin after the evacuation and mix with the population therein. To account for the increased human interactions associated with evacuating to shared living spaces^7^, we additionally assume the transmission rates in the origin and destination are elevated during a period that spans the evacuation process. Specifically, the transmission rate in the origin is increased by 20% starting from 3 days prior to the evacuation until 3 days after the return of evacuees. The transmission rate in the destination is increased by 20% during the evacuation.

In model simulations, we fixed the following parameters in Eqs. (1)-(5): total population *N*_1_ = *N*_2_ *=* 10^6^; reporting rate *α =* 0.1; relative transmissibility *μ =* 0.64; latency period *Z* = 4 days; infectious period *D* = 4 days. Denote the daily reported cases in the origin and destination as *case_i_*. To initiate model simulations, we set 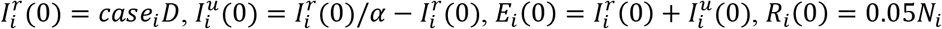 and 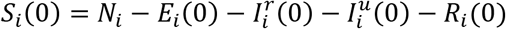. Model simulations were generated for the following stages after day 0: 14 days of local transmission, 3 days of pre-evacuation (with elevated transmission rate in the origin), *T_eva_* days of evacuation (with elevated transmission rates in both the origin and destination counties), 3 days of post-evacuation (with elevated transmission rate in the origin), and 28 days of additional post-evacuation simulation.

We varied six parameters to generate a large number of parameter combinations for use in model simulation: *β*_1_*, β*_2_ *=* 0.1, 0.2,…,0.8; *T_eva_ =* 3, 4,…,10 days; *p_eva_ =* 0.1,0.2,…,0.8; *case*_1_*, case*_2_ *=* 50,100,…, 400. In total, 8^6^ = 262,144 parameter combinations were simulated. For comparison, we also ran a simulation without evacuation for each parameter combination and computed the percentage change of total cases in the origin and destination counties attributed to evacuation. We selected the top 10% of combinations that lead to the lowest percentage increase (or highest percentage reduction) of reported cases in the origin county, the destination county and both counties combined, and inspected the marginal distributions of those six parameters.

### Simulating COVID-19 transmission in the US

Following the two-county model analysis, we conducted in-depth COVID-19 simulations using a nation-wide metapopulation SEIR model representing all 3,142 US counties^9^. In this model, disease transmission in each county follows SEIR dynamics, but is also influenced by movement to and from other counties. We consider two types of movement: daily work commuting and random movement. To simulate movement prior to March 15, 2020, we use information on county-to-county work commuting that is publicly available from the US Census Bureau. After March 15, the census survey data are no longer representative due to changes of mobility behavior in response to COVID-19 control measures.

Therefore, in simulating movement after March 15, 2020, we use estimates of the reduction of inter-county visitors to points of interest (POI) (e.g., restaurants, stores, etc.) to inform the decline of inter-county movement on a county-by-county basis. We generated these estimates using data from SafeGraph^56^. We further assume the number of random visitors between two counties is proportional to the average number of commuters between them. As population present in each county is different during daytime and nighttime, we model the transmission dynamics of COVID-19 separately for these two time periods. The model equations are presented in the supplementary information. Similar models have been used to simulate COVID-19 transmission in China^57^ and influenza transmission in the United States^58,59^. The local effective reproductive number is derived as *R_e_ = βiD[α + μ*(1 − *α*)*]S_i_/N_i_*. To account for reporting delays in COVID-19 case and death observations, we mapped simulated documented infections to confirmed cases using a separate observational delay model fitted to the US case data^59^.

We calibrated the transmission model against county-level case and death data reported from February 21, 2020 through July 23, 2020^56,60^, which produced an estimate of model parameters and state variables. We then ran simulations representing the following stages: 3 days of pre-evacuation, 7 days of evacuation, 3 days of post-evacuation and 14 days after post-evacuation. For comparison, we also ran simulations without evacuation and increase of transmission rates in origin and destination counties. An ensemble of 100 trajectories were generated to represent the uncertainty arising from different initial conditions and stochastic dynamics.

In the modeled hurricane evacuation, *V_ji_* evacuees travel from origin *i* to destination *j* and mix with the local population for *T_eva_ =* 7 days, before returning to origin *i*. As for the two-county model, we increased the transmission rate in origin counties by 20% during the 3-day pre-evacuation, 7-day evacuation and 3-day post-evacuation. Three scenarios in which the transmission rate in destination counties is increased by 0%, 10% and 20% were simulated to compare different effects of hosting evacuees on local disease transmission.

### The greedy algorithm to optimize evacuation

We developed a greedy optimization algorithm aimed at minimizing total excess COVID-19 cases by strategically assigning evacuees to optimal destination counties. In the evacuation optimization, we assume that a fraction *p* of evacuees from an origin to a destination won’t change their evacuation plans (for reasons such as personal connections at the destination or budgetary limitations) and the capacity of accepting evacuees for each destination *j* is *C_j_*. Denote the baseline evacuation matrix as ***V***, where *V_ji_* represents the number of evacuees from origin *i* to destination *j*. The optimization objective is to assign the rest of evacuees (i.e., (1 − *p*) *× ∑_j_ V_ji_* from origin *i*) to destinations in an optimal way that minimizes the total infections in both origin and destination counties. Finding the exact solution to this combinatorial optimization problem is computationally challenging due to the large number of options.

In this study, we use a practically feasible greedy optimization approach that prioritizes moving the unassigned evacuees to destinations with low *R_e_*. Specifically, we start from the evacuation matrix *p****V*** that represents evacuees assigned a destination. In each step of greedy search, we run a series of simulations, each one filling the available evacuee slots in the destination with the lowest *R_e_* from one of the origin counties. We select the origin county that generates the minimum number of reported cases and assign them to the destination county. We repeat this greedy search for each successive destination county until all evacuees are assigned a destination. The pseudo-code for this greedy algorithm is provided in the supplementary information. In this study, we assume 10% of evacuees from an origin to a destination in the baseline evacuation matrix ***V*** cannot be reallocated (i.e., *p =* 0.1), and the capacity of each destination is 120% of the evacuees in the baseline scenario ***V*** (i.e., *C_j_* = 1.2 ∑*_i_ V_ji_*).

## Supplementary Information

### Transmission model for 3,142 US counties

We formulate COVID-19 transmission as a discrete Markov process during both day and night. Daytime transmission lasts for *dt*_1_ days and the nighttime transmission *dt*_2_ days (*dt*_1_ + *dt*_2_ *=* 1). Here, we assume daytime transmission lasts for 8 hours and nighttime transmission lasts for 16 hours, i.e., *dt*_1_ = 1/3 day and *dt*_2_ = 2/3 day. The transmission dynamics are depicted by the following equations.

Daytime transmission:

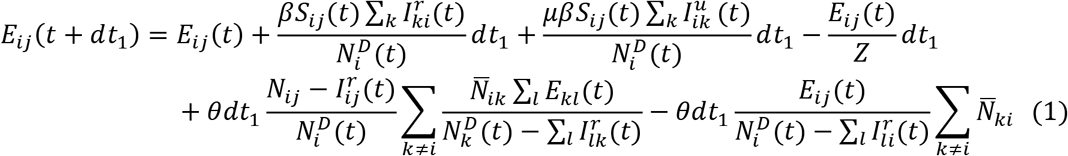

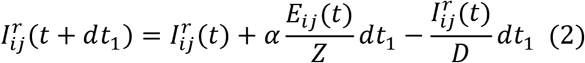

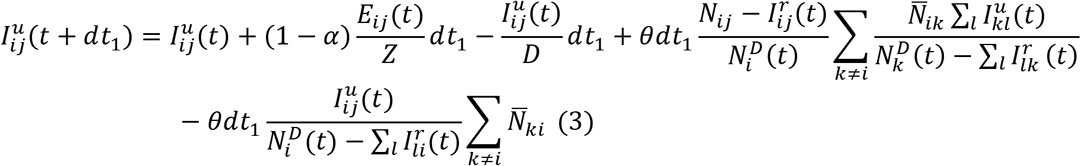

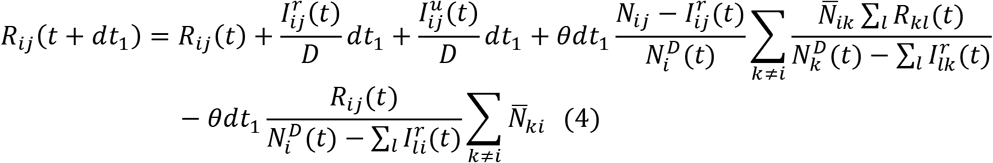

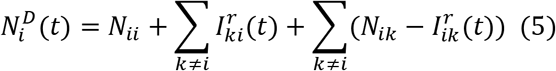

Nighttime transmission:

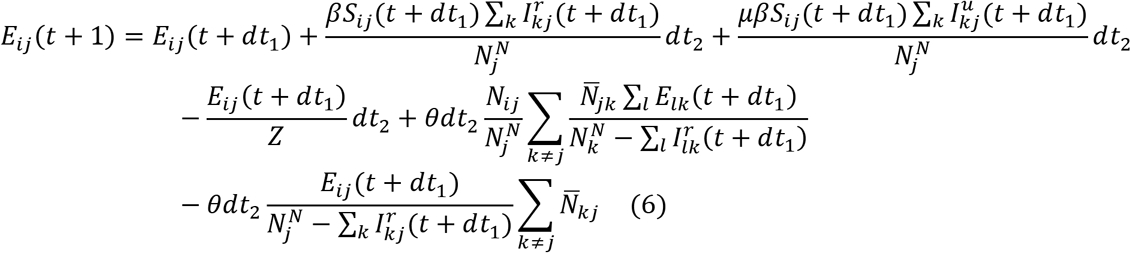

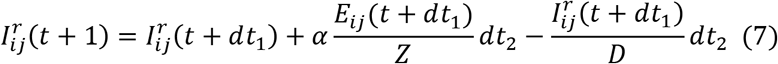

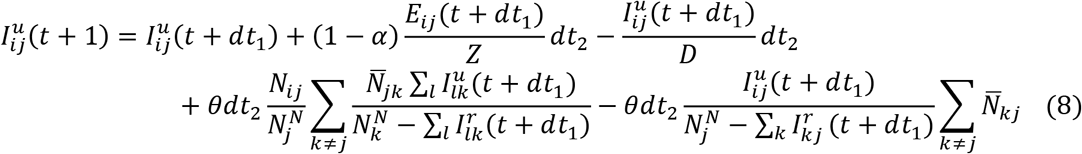

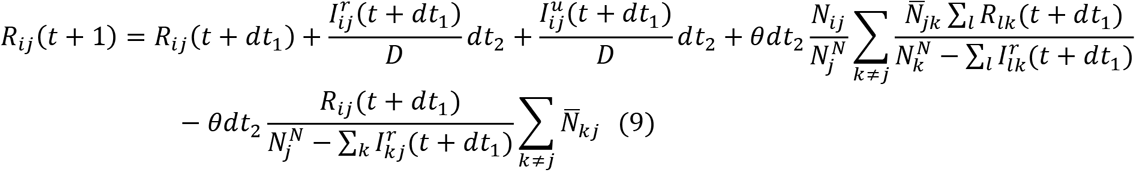

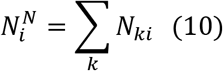

Here, *S_ij_, E_ij_*, 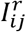, 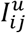 *R_ij_* and *N_ij_* are the susceptible, exposed, reported infected, unreported infected, recovered and total populations in the subpopulation commuting from county *j* to county *i* (*i → j*), where 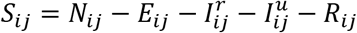; *β* is the transmission rate of reported infections; *μ* is the relative transmissibility of unreported infections; *Z* is the average latency period (from infection to contagiousness); *D* is the average duration of contagiousness; *α* is the fraction of documented infections; *θ* is a multiplicative factor adjusting random movement; 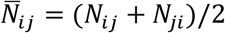 is the average number of commuters between counties *i* and *j*; and 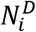 and 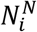 are the daytime and nighttime populations of county *i*.

### The pseudo-code for the greedy optimization algorithm

Input:

Origin *i =* 1,2,…,*n*

Destination *j* = 1,2,…*.,m*, where *R_e_*(1) ≥ *R_e_*(2) *≥…*≥ *R_e_*(*m*)

Evacuation matrix ***V*** *= {V_ji_}, V_ji_* is the number of evacuees from origin *i* to destination *j* in the baseline scenario

Capacity of evacuees that can be accommodated by each destination: *C_j_*

The fraction of evacuees that can’t be reallocated for each origin-destination pair: *p*

Variables:

*u_i_*:: the current number of evacuees in origin *i* that could be reallocated to different counties

*ν:* the currently available destination county with lowest *R_e_*.

***M***: the current evacuation matrix, *M_νi_* is the number of evacuees assigned from origin *i* to destination *ν*.

Initial conditions:

*ν = m*

*u_i_ =* (1*−p*)*∑_j_V_ji_*

***M*** *= p****V***

Algorithm:

While max(*u_i_*) > 0

For *i =* 1 to *n*

***M****^i^ =* ***M***: reallocating from origin *i* to *ν*

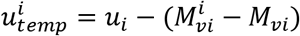

Run projection using *M^i^*

*Inf_i_*: total infection in all origin and destination counties

End

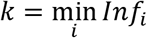

***M*** = ***M****^k^*

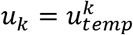

If ∑*j* ***M****_νj_* == *C_ν_*

End

End

Output ***M*** as the optimized evacuation matrix.

**Fig. S1.**
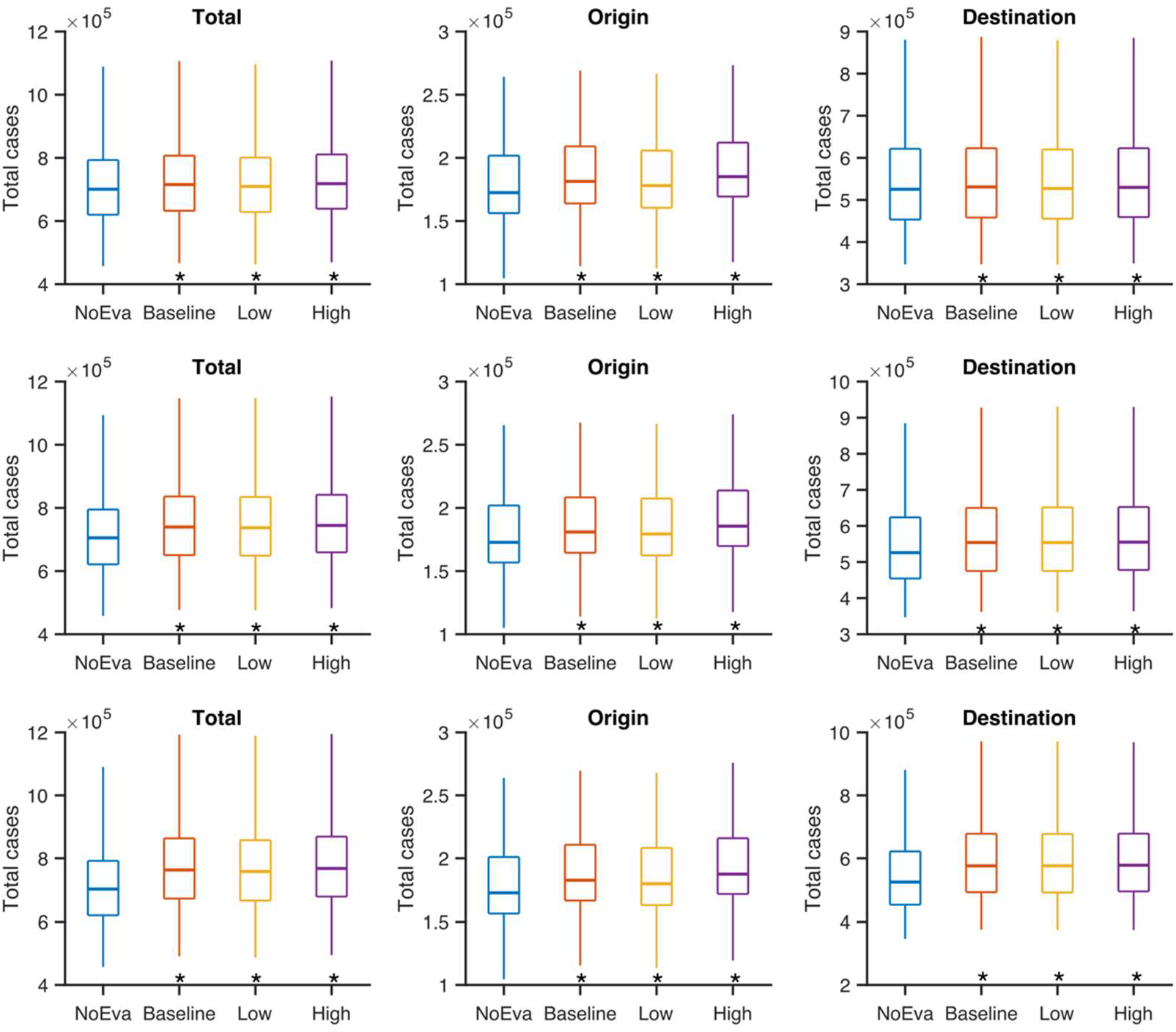
Comparison of total cases in the origin and destination counties combined (left column), the origin counties only (middle column) and destination counties only (right column) for the no-evacuation, baseline, low and high evacuation scenarios. Simulations were performed for three settings: no increase (top row), 10% increase (middle row) and 20% increase (bottom row) of transmission rates in destination counties. Box plots show the median and interquartile and whiskers show the 95% CIs. Asterisks indicate that excess cases are significantly higher than the no-evacuation scenario (Wilcoxon signed rank test, *p <* 10^−5^).

**Fig. S2.**
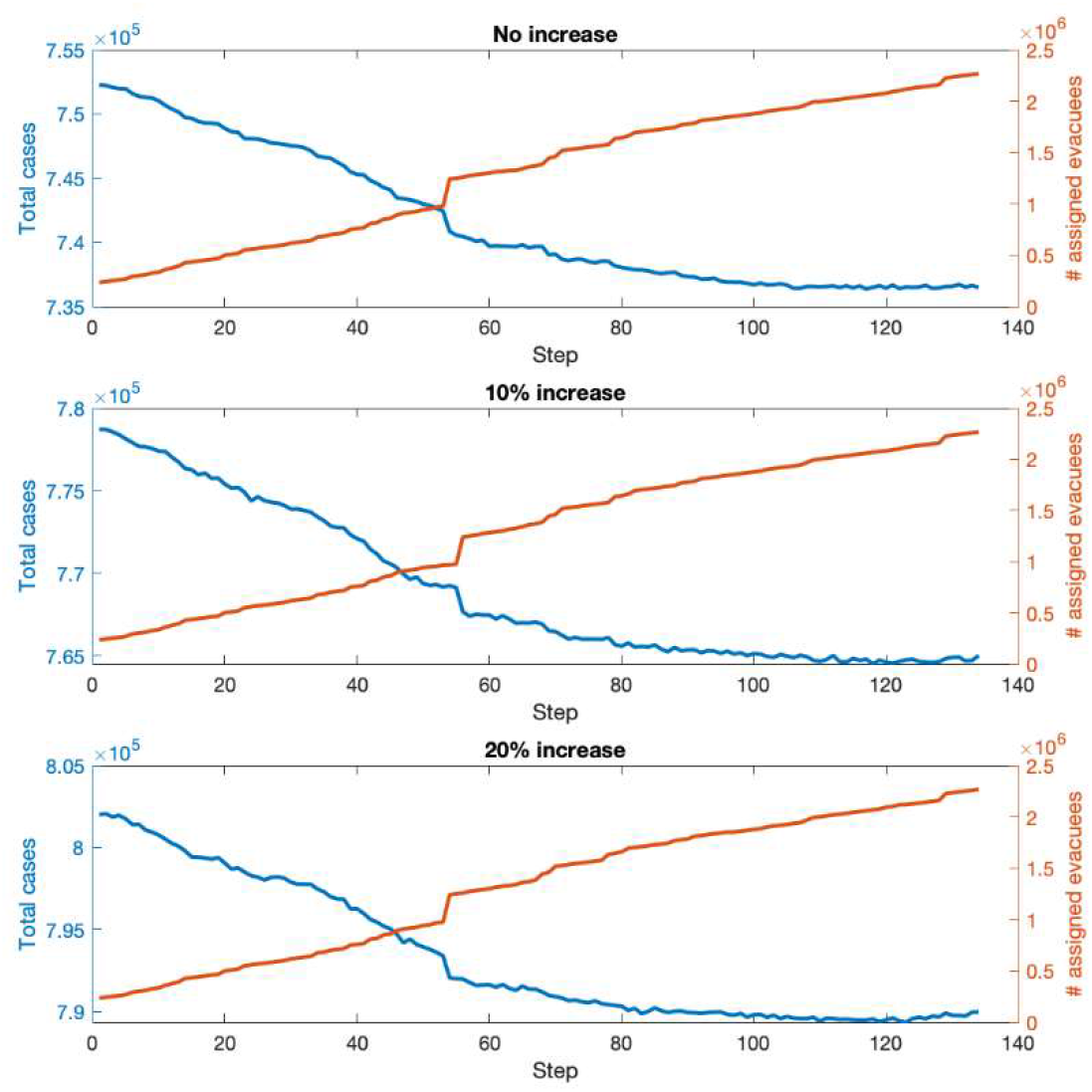
Evolution of the total cases (blue lines) and the number of assigned evacuees (red lines) in the greedy algorithm. Results are shown for the settings with no increase, 10% increase and 20% increase of transmission rates in destination counties. The optimization starts from an evacuation matrix 0.1 × ***V***, where ***V*** represents the evacuation matrix in the baseline scenario.

**Table S1.** The median number of total cases in origin and destination counties for different evacuation scenarios (no evacuation, baseline, low and high) and levels of elevated transmission rates (*R_e_*) in destination counties (no change, 10% increase, 20% increase). In the no evacuation scenario (*R_e_*) in destination counties is not increased. Note that the median excess cases shown in Table 1 is not necessarily the difference of the median total cases between the evacuation and non-evacuation scenarios shown in Table S1. That is, *median{totalcase_eva_*(*i*) *− totalcase_noeva_*(*i*)) ≠ *median{totalcase_eva_*(*i*)) *- median{totalcase_noeva_*(*i*)), where *totalcase_eva_*(*i*) and *totalcase_noeva_*(*i*) are the total numbers of cases for evacuation and non-evacuation scenarios in the *i*th simulation.

| | 0% $R_e$ increase in destination | | 10% $R_e$ increase in destination | | 20% $R_e$ increase in destination | |
| --- | --- | --- | --- | --- | --- | --- |
|  | Origin cases | Destination cases | Origin cases | Destination cases | Origin cases | Destination cases |
| **No Evacuation** | 172,400 | 525,320 | 172,692 | 526,044 | 172,562 | 524,467 |
| **Baseline evacuation** | 181,322 | 530,706 | 180,901 | 553,971 | 182,590 | 575,891 |
| **High $R_e$**<br>**evacuation** | 185,100 | 529,781 | 185,527 | 555,062 | 187,431 | 577,885 |
| **Low $R_e$**<br>**evacuation** | 178,029 | 527,180 | 179,317 | 553,875 | 179,788 | 576,043 |

**Table S2.**
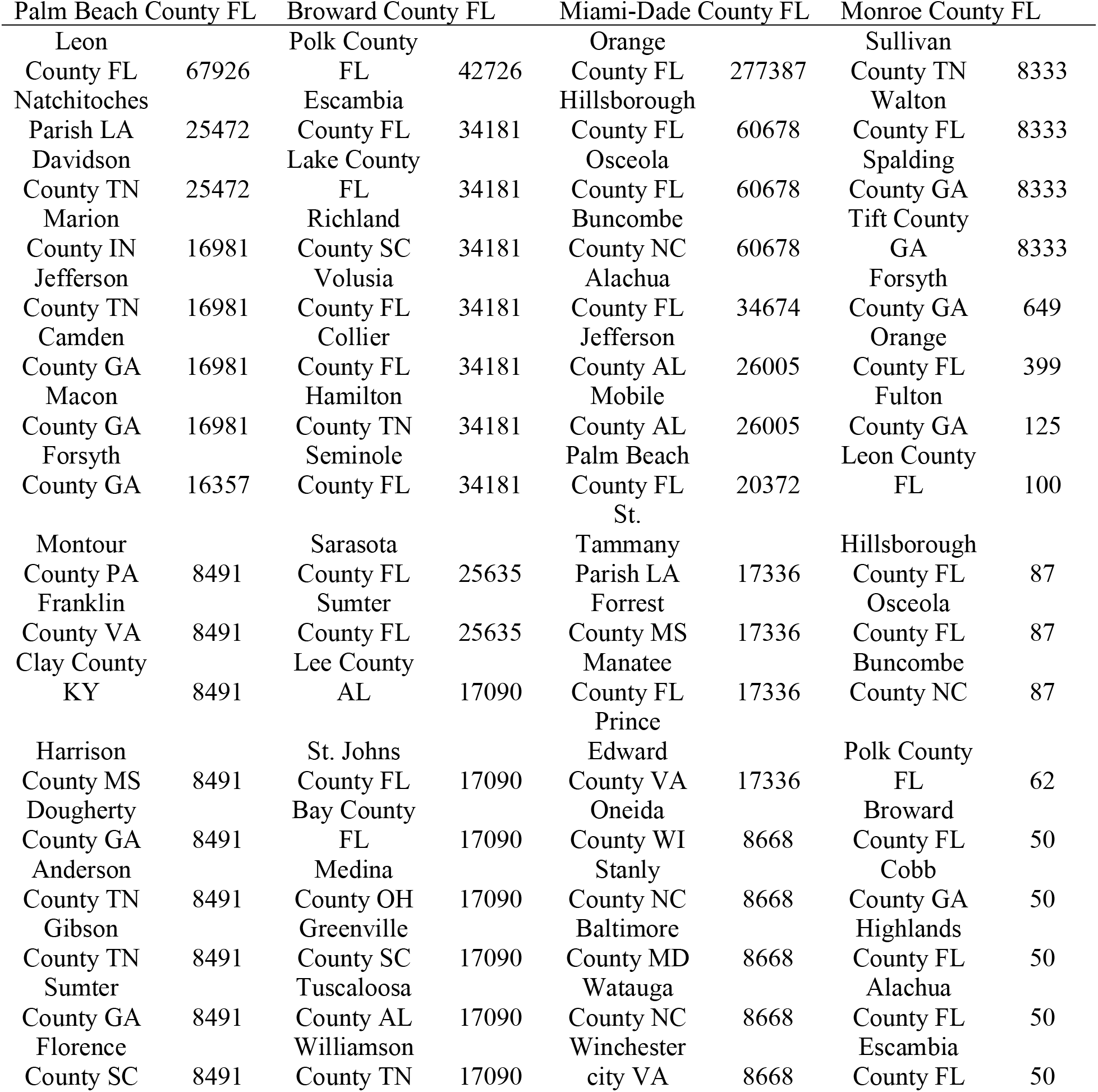
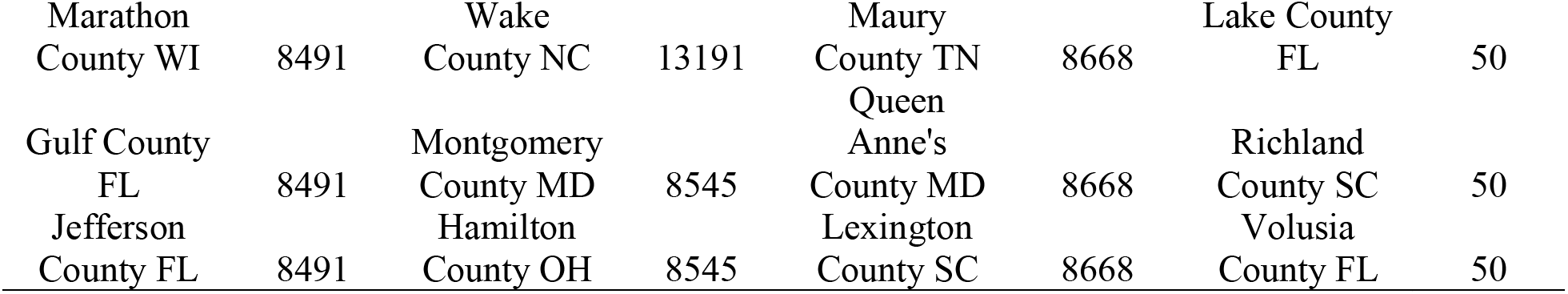
The optimized evacuation plan (no increase of transmission rates in destination counties). We show the top 20 destinations for each origin county.

**Table S3.** The optimized evacuation plan (10% increase of transmission rates in destination counties). We show the top 20 destinations for each origin county.

| Palm Beach County<br>FL |  | Broward County FL |  | Miami-Dade County FL |  | Monroe County FL |  |
| --- | --- | --- | --- | --- | --- | --- | --- |
| Leon<br>County FL | 67926 | Polk County<br>FL | 42726 | Orange<br>County FL | 277387 | St. Johns<br>County FL | 8969 |
| Jefferson<br>County AL | 25472 | Escambia<br>County FL | 34181 | Hillsborough<br>County FL | 60678 | Charleston<br>County SC | 8333 |
| Mobile<br>County AL | 25472 | Lake<br>County FL | 34181 | Osceola<br>County FL | 60678 | Walton<br>County FL | 8333 |
| Marion<br>County IN | 16981 | Richland<br>County SC | 34181 | Buncombe<br>County NC | 60678 | Oakland<br>County MI | 8333 |
| Macon<br>County<br>GA | 16981 | Volusia<br>County FL | 34181 | Alachua<br>County FL | 34674 | Orange<br>County FL | 399 |
| Lee<br>County AL | 16981 | Collier<br>County FL | 34181 | Natchitoches<br>Parish LA | 26005 | Fulton<br>County GA | 125 |
| Jefferson<br>County TN | 11527 | Hamilton<br>County TN | 34181 | Davidson<br>County TN | 26005 | Leon County<br>FL | 100 |
| Montour<br>County PA | 8491 | Seminole<br>County FL | 34181 | Palm Beach<br>County FL | 20372 | Hillsborough<br>County FL | 87 |
| Franklin<br>County<br>VA | 8491 | Sarasota<br>County FL | 25635 | Camden<br>County GA | 17336 | Osceola<br>County FL | 87 |
| Clay<br>County<br>KY | 8491 | Sumter<br>County FL | 25635 | St.<br>Tammany<br>Parish LA | 17336 | Buncombe<br>County NC | 87 |
| Harrison<br>County<br>MS | 8491 | Bay County<br>FL | 17090 | Forrest<br>County MS | 17336 | Polk County<br>FL | 62 |
| Dougherty<br>County<br>GA | 8491 | Manatee<br>County FL | 17090 | Forsyth<br>County GA | 17336 | Broward<br>County FL | 50 |
| Anderson<br>County TN | 8491 | Medina<br>County OH | 17090 | Wake<br>County NC | 17336 | Cobb<br>County GA | 50 |
| Gibson<br>County TN | 8491 | Prince<br>Edward<br>County VA | 17090 | Tuscaloosa<br>County AL | 17336 | Highlands<br>County FL | 50 |
| Sumter<br>County<br>GA | 8491 | Greenville<br>County SC | 17090 | Williamson<br>County TN | 17336 | Alachua<br>County FL | 50 |
| Florence<br>County SC | 8491 | Martin<br>County FL | 8545 | Gulf County<br>FL | 8668 | Escambia<br>County FL | 50 |
| Marathon<br>County WI | 8491 | Baker<br>County FL | 8545 | Jefferson<br>County FL | 8668 | Lake County<br>FL | 50 |
| Towns<br>County<br>GA | 8491 | Ben Hill<br>County GA | 8545 | Greene<br>County TN | 8668 | Richland<br>County SC | 50 |
| Frederick<br>County<br>VA | 8491 | Santa Cruz<br>County CA | 8545 | Jackson<br>County GA | 8668 | Volusia<br>County FL | 50 |
| Pitt County<br>NC | 8491 | Rockingham<br>County VA | 8545 | Madison<br>County MS | 8668 | Collier<br>County FL | 50 |

**Table S4.** The optimized evacuation plan (20% increase of transmission rates in destination counties). We show the top 20 destinations for each origin county.

| Palm Beach County<br>FL |  | Broward County FL |  | Miami-Dade County FL |  | Monroe County FL |  |
| --- | --- | --- | --- | --- | --- | --- | --- |
| Leon<br>County FL | 67926 | Polk<br>County FL | 42726 | Orange<br>County FL | 277387 | Dodge<br>County GA | 8333 |
| Davidson<br>County TN | 25472 | Escambia<br>County FL | 34181 | Hillsborough<br>County FL | 60678 | St. Lucie<br>County FL | 8333 |
| Jefferson<br>County AL | 25472 | Lake<br>County FL | 34181 | Osceola<br>County FL | 60678 | Erie County<br>NY | 8333 |
| Marion<br>County IN | 16981 | Richland<br>County SC | 34181 | Buncombe<br>County NC | 60678 | Maricopa<br>County AZ | 8333 |
| Jefferson<br>County TN | 16981 | Volusia<br>County FL | 34181 | Alachua<br>County FL | 34674 | Lexington<br>County SC | 637 |
| Wake<br>County NC | 16981 | Collier<br>County FL | 34181 | Natchitoches<br>Parish LA | 26005 | Orange<br>County FL | 399 |
| Forsyth<br>County<br>GA | 12560 | Hamilton<br>County TN | 34181 | Mobile<br>County AL | 26005 | Fulton<br>County GA | 125 |
| Montour<br>County PA | 8491 | Seminole<br>County FL | 34181 | Palm Beach<br>County FL | 20372 | Leon County<br>FL | 100 |
| Franklin<br>County<br>VA | 8491 | Sarasota<br>County FL | 25635 | Camden<br>County GA | 17336 | Hillsborough<br>County FL | 87 |
| Clay<br>County<br>KY | 8491 | Sumter<br>County FL | 25635 | Macon<br>County GA | 17336 | Osceola<br>County FL | 87 |
| Harrison<br>County<br>MS | 8491 | St. Johns<br>County FL | 17090 | St.<br>Tammany<br>Parish LA | 17336 | Buncombe<br>County NC | 87 |
| Dougherty<br>County<br>GA | 8491 | Bay County<br>FL | 17090 | Forrest<br>County MS | 17336 | Polk County<br>FL | 62 |
| Anderson<br>County TN | 8491 | Manatee<br>County FL | 17090 | Lee County<br>AL | 17336 | Broward<br>County FL | 50 |
| Gibson<br>County TN | 8491 | Medina<br>County OH | 17090 | Tuscaloosa<br>County AL | 17336 | Cobb<br>County GA | 50 |
| Sumter<br>County<br>GA | 8491 | Prince<br>Edward<br>County VA | 17090 | Gulf County<br>FL | 8668 | Highlands<br>County FL | 50 |
| Marathon<br>County WI | 8491 | Greenville<br>County SC | 17090 | Jefferson<br>County FL | 8668 | Alachua<br>County FL | 50 |
| Towns<br>County<br>GA | 8491 | Williamson<br>County TN | 17090 | Greene<br>County TN | 8668 | Escambia<br>County FL | 50 |
| Frederick<br>County<br>VA | 8491 | Winston<br>County AL | 8545 | Jackson<br>County GA | 8668 | Lake County<br>FL | 50 |
| Pitt County<br>NC | 8491 | Escambia<br>County AL | 8545 | Madison<br>County MS | 8668 | Richland<br>County SC | 50 |
| Habersham<br>County<br>GA | 8491 | Robeson<br>County NC | 8545 | Oneida<br>County WI | 8668 | Volusia<br>County FL | 50 |

